# Racial/ethnic disparities in exposure to COVID-19, susceptibility to COVID-19 and access to health care – findings from a U.S. national cohort

**DOI:** 10.1101/2022.01.11.22269101

**Authors:** McKaylee M Robertson, Meghana Shamsunder, Ellen Brazier, Mekhala Mantravadi, Madhura S. Rane, Drew A. Westmoreland, Angela M. Parcesepe, Rebecca Zimba, Andrew R. Maroko, Sarah G. Kulkarni, Christian Grov, Denis Nash

**Affiliations:** Institute for Implementation Science in Population Health (ISPH); 55 W 125th St, 6th Floor, New York, NY 10027, USA; Graduate School of Public Health and Health Policy; 55 W 125th St, New York, NY 10027, USA; Gillings School of Public Health, University of North Carolina, 427 Rosenau Hall, CB #7445, Chapel Hill, NC 27599-7445, USA; Carolina Population Center, University of North Carolina at Chapel Hill, Chapel Hill, NC, USA

**Keywords:** COVID-19, healthcare disparities

## Abstract

We examined the influence of racial/ethnic differences in socioeconomic position on COVID-19 seroconversion and hospitalization within a community-based prospective cohort enrolled in March 2020 and followed through October 2021 (N=6740). The ability to social distance as a measure of exposure to COVID-19, susceptibility to COVID-19 complications, and access to healthcare varied by race/ethnicity with non-white participants having more exposure risk and more difficulty with healthcare access than white participants. Participants with more (versus less) exposure had greater odds of seroconversion (aOR:1.64, 95% Confidence Interval [CI] 1.18-2.29). Participants with more susceptibility and more barriers to healthcare had greater odds of hospitalization (respective aOR:2.36; 1.90-2.96 and 2.31; 1.69-2.68). Race/ethnicity positively modified the association between susceptibility and hospitalization (aOR_non-White_:2.79, 2.06-3.78). Findings may explain the disproportionate burden of COVID-19 infections and complications among Hispanic and non-Hispanic Black persons. Primary and secondary prevention efforts should address disparities in exposure, COVID-19 vaccination, and treatment.

## INTRODUCTION

Researchers have identified underlying medical conditions, comorbidities, older age, and male sex as biological vulnerabilities for more severe Coronavirus Disease 2019 (COVID-19) outcomes.(1,2) Evidence also suggests a disproportionate burden of COVID-19 infection, hospitalization, and death among Hispanic and Black Non Hispanic (NH) individuals in the United States (U.S.).(3–6) Early in the pandemic (March 2020) the Centers for Disease Control and Prevention (CDC) reported that twice as many Black individuals were hospitalized with COVID-19 than are proportionally represented in the U.S.(3) Long-standing health and social inequities likely contribute to disparities in COVID-19 morbidity and mortality.(7–9)

Public health interventions and policies with the potential to improve health may inadvertently amplify existing health disparities.(7) Prevention efforts, such as social distancing or work from home policies, may not benefit racial and ethnic minority groups as much as non-minority and more advantaged groups, given they are disproportionately represented in essential work settings and/or more likely to live in multi-generational households.(7,8,10) Less access to and utilization of healthcare also result in worse COVID-19 outcomes among racial and ethnic minority groups since later care presentation provides less opportunity to benefit from various treatment modalities.(8,11) Barriers to healthcare access may also increase the likelihood of staying at home longer while infectious. Blumenshine et al., proposed a pandemic disease model in which differences in socioeconomic position (SEP) contribute to disparities in **exposure, susceptibility** to illness, if exposed, and **access** to treatment before disease development.(12) In the Blumenshine model, disparities in exposure, susceptibility, and access may lead to unequal levels of infection, morbidity and mortality during a pandemic. To avoid exacerbating existing disparities, effective public health interventions and pandemic guidelines need to anticipate and mitigate the contribution of SEP to racial/ethnic disparities.(9,12,13)

Our objective was to examine the influence of racial and ethnic differences in SEP on COVID-19 outcomes within a large U.S. national cohort that was enrolled during the spring of 2020, the early phase of the COVID-19 pandemic. Using the Blumenshine model as a framework, we created three indices to assess SEP: a) the ability to social distance as a measure of exposure to COVID-19, b) susceptibility to COVID-19 complications, and c) access to healthcare. We examined the relationship between each SEP index with COVID-19 outcomes (COVID-19 hospitalization or seroconversion), and we assessed race/ethnicity as a potential effect measure modifier (EMM) of the relationship between each SEP index and COVID-19 outcome.

## METHODS

### Data Source and Population

The CHASING COVID Cohort study is a geographically and socio-demographically diverse sample of adults (18 and older), residing in the U.S. or U.S. territories and enrolled into a prospective cohort study during the emergence of the U.S. COVID-19 pandemic.(14) Study participants were recruited from March 28, 2020 to April 20, 2020 via social media (e.g., Facebook) or via referral. Details of cohort recruitment and follow-up been described elsewhere.(14)

### Research Ethics Approval

The study protocol was approved by the Institutional Review Board at the City University of New York (CUNY) Graduate School for Public Health and Health Policy.

#### Variable Definitions

##### Race/Ethnicity

Respondents were asked: “Are you Hispanic, Latino/a, or Spanish origin?” and “Which of these groups would you say best represents your race?” Participants were then categorized as Hispanic, Black Non-Hispanic (NH), Asian/Pacific Islander (API) NH, (which included American Indian or Alaskan Native), White NH and Other.(15)

##### Exposures: COVID-19 Exposure, COVID-19 Susceptibility, and Healthcare Access

We created three indices related to: 1) COVID-19 exposure, 2) susceptibility and 3) access to health care.(9) The indices and assessment items were taken from a national survey that explored the experience of adults during the H1N1 pandemic. ^(9,16)^ Each index was a summative score, where a higher risk response was given a value of 1, and a lower or no risk response was given a value of 0. Therefore, a higher value would indicate a greater exposure risk, greater susceptibility, and greater difficulty with access to care and treatment.

First, as the measure of **COVID-19 exposure**, we included structural and work-related items that contributed to the ability to social distance. The structural items included: living in an urban area, living in a multi-unit dwelling (e.g., apartment building), and the ability to avoid public transportation. The work-related items included essential worker status and whether respondents were able to stay home from work or work from home, if needed. Specifically, respondents were asked to indicate “yes”, “no” or “not applicable” to the following: (1) I am able to work at home; (2) If I do not go to work because I am ill, I will not get paid for the time I am at home; (3) I have sick leave at my job if I need to use it; (4) I could lose my job or business if I am not able to go into work; (5) My job can only be done in my workplace. Respondents who did not work were considered not at risk for the work-related items (i.e., a score of zero). Essential worker status was defined as having been involved in healthcare or other essential work (e.g., first responders) in the two weeks prior to the survey.(14)

Second, as the measure of **COVID-19 susceptibility**, we included conditions or behaviors that CDC has identified as increasing the risk of COVID-19 complications given SARS-CoV-2 infection: aged 60 years or older, daily smoking, and underlying chronic conditions (chronic lung disease, current asthma, type 2 diabetes, kidney disease, immunocompromised condition, or HIV positive). Finally, as the measure of **healthcare access**, we included factors that affect medical care access: no primary care doctor, concerns about the costs of healthcare, concerns about seeing a doctor due to immigration status, or no health care coverage/insurance.

We dichotomized each index as less than or equal to the median value for statistical models: more versus less exposure risk, more versus less susceptible, and more versus less difficulty with access to care. Exposure data came from baseline recruitment surveys.

##### COVID-19 Outcomes: Hospitalization and Seroconversion

We examined the association of exposure, susceptibility, or access to care with two COVID-19 outcomes: COVID-19 hospitalization and seroconversion. We defined **COVID-19 hospitalization** as a self-report of hospitalization for any COVID-19-like symptoms from baseline through the eighth follow-up assessment (V0-V8, March 2020-October 2021). “Since you completed your last survey on DD/MM/YYYY, were you hospitalized for any of these symptoms?” The outcome was dichotomized as “yes” and “no” with those who reported “do not know” or “not sure” classified as “no.”

The procedure for at-home specimen collection for serologic testing has been described.(17) Briefly, participants were invited to participate in serologic testing using an at-home self-collected dried blood spot (DBS) specimen collection kit during May-August 2020 (Period 1) and November 2020-January 2021 (Period 2). All DBS specimens were tested by the study laboratory for total antibodies (Total Ab) using the Bio-Rad Platelia test for IgA, IgM, and IgG which targets the SARS-CoV-2 nucleocapsid protein. (18) Among those individuals with two Total Ab tests, an **observed seroconversion** was defined as a negative Total Ab test in Period 1 followed by a positive Total Ab test in Period 2.

##### Confounders

Age, gender, presence of children in the household, income, education, and loss of income or employment were treated as confounders of the hypothesized exposure-outcome relationships.

##### Statistical Analysis

We used descriptive statistics to examine participant demographics and SEP indices reflecting COVID-19 exposure, susceptibility, and access to healthcare stratified by race/ethnicity. We assessed differences between groups using the Chi-Square or Kruskal Wallis test as appropriate.

We used a logistic regression model to assess the association between each SEP index and outcomes of interest: COVID-19-hospitalization or seroconversion. We separately modelled each exposure-outcome relationship. When COVID-19 exposure or access to health care was the explanatory index of interest, the models were adjusted for confounders and the exposure index. When susceptibility was the explanatory index of interest, we adjusted for all confounders except age, as age was used to create the susceptibility summative score.

We assessed whether the effect of each index on COVID-19 outcomes was modified by race/ethnicity. We assessed EMM on the additive scale and present the relative excess risk due to interaction (RERI).(19,20) As EMM on the additive scale indicates whether the effect of an exposure is different in one subpopulation relative to another, it is considered useful for for identifying specific populations for whom public health interventions will have the most impact.(19,20) Because of small cells, we collapsed the race variable to White NH versus non-White for assessment of EMM.

We conducted logistic regression models with SAS (V9.4) and generated confidence intervals for the RERI using the spreadsheet tool published by Knol et al.(19)

## RESULTS

This analysis utilized data from 6,740 individuals enrolled into prospective follow-up for analyses assessing the hospitalization outcome reported through October 20, 2021. Among the full cohort 19% (N=1,308) identified as Hispanic ethnicity, 13% (N=899) as Black NH, 7% (N=465) as API NH, 57% (N=3,846) as White NH, and 3% (N=222) as other NH race (Table 1). Hispanics, Black NH, or API NH were younger on average than White NH (mean age in years [Standard Deviation]: 35 [13], 35 [13], 33 [12] versus 45 [16], respectively). More than half of the cohort was women (52%). Over half of the cohort (57%) had a college-level education, and the proportion with a college-education was highest among API NH (69%) and lowest among Black NH participants (33%).

**Table 1.** Demographic and Socioeconomic Characteristics of Chasing COVID Cohort (C3) Participants Stratified by Race and Ethnicity - United States, March 28 - April 20, 2020.

|  | Total | Hispanic | Black NH | Asian\Pacific<br>Islander NH | White NH | Other NH | P-value |
| --- | --- | --- | --- | --- | --- | --- | --- |
| **Total, n (%\*)** | 6,740 (100.00) | 1,308 (19.41) | 899 (13.33) | 465 (6.90) | 3,846 (57.06) | 222 (3.30) |  |
| **Age** |  |  |  |  |  |  | < 0.001 |
| Mean (SD) | 40.61 (15.28) | 35.19 (13.33) | 35.31 (12.80) | 32.73 (11.95) | 44.64 (15.54) | 40.74 (14.06) |  |
| Median (IQR) | 37 (29, 51) | 33 (25, 42) | 32 (26, 42) | 30 (24, 39) | 42 (32, 57) | 39 (29, 49) |  |
| **Age Category, n (%)** |  |  |  |  |  |  | < 0.001 |
| 18-29 | 1,802 (26.75) | 496 (37.92) | 357 (39.71) | 217 (46.67) | 673 (17.5) | 60 (27.03) |  |
| 30-39 | 1,956 (29.02) | 424 (32.42) | 275 (30.59) | 139 (29.89) | 1,066 (27.72) | 52 (23.42) |  |
| 40-49 | 1,163 (17.26) | 214 (16.36) | 123 (13.68) | 68 (14.62) | 701 (18.23) | 57 (25.68) |  |
| 50-59 | 792 (11.75) | 99 (7.57) | 90 (10.01) | 19 (4.09) | 559 (14.53) | 25 (11.26) |  |
| 60+ | 1,027 (15.22) | 75 (5.73) | 54 (6.01) | 22 (4.73) | 847 (22.02) | 28 (12.61) |  |
| **Gender, n (%)** |  |  |  |  |  |  | < 0.001 |
| Male | 3,043 (45.15) | 568 (43.43) | 411 (45.72) | 195 (41.94) | 1,762 (45.81) | 107 (48.2) |  |
| Female | 3,526 (52.31) | 718 (54.89) | 468 (52.06) | 260 (55.91) | 1,983 (51.56) | 97 (43.69) |  |
| Non-Binary | 171 (2.54) | 22 (1.68) | 20 (2.22) | 10 (2.15) | 101 (2.63) | 18 (8.11) |  |
| **Education, n (%)** |  |  |  |  |  |  | < 0.001 |
| Less than 12th grade | 123 (1.82) | 34 (2.6) | 25 (2.78) | 9 (1.94) | 54 (1.4) | 1 (0.45) |  |
| 12th grade/GED | 875 (12.98) | 282 (21.56) | 191 (21.25) | 36 (7.74) | 330 (8.58) | 36 (16.22) |  |
| Some college (1-3 years) | 1,889 (28.03) | 436 (33.33) | 385 (42.83) | 100 (21.51) | 894 (23.24) | 74 (33.33) |  |
| College (4 or more years) | 3,853 (57.17) | 556 (42.51) | 298 (33.15) | 320 (68.82) | 2,568 (66.77) | 111 (50.00) |  |
| **Employment Status, n (%)** |  |  |  |  |  |  | < 0.001 |
| Employed | 4,247 (63.01) | 811 (62) | 587 (65.29) | 267 (57.42) | 2,443 (63.52) | 139 (62.61) |  |
| Out of Work | 830 (12.31) | 206 (15.75) | 131 (14.57) | 55 (11.83) | 402 (10.45) | 36 (16.22) |  |
| Other | 1,663 (24.67) | 291 (22.25) | 181 (20.13) | 143 (30.75) | 1,001 (26.03) | 47 (21.17) |  |
| **Income, n (%)** |  |  |  |  |  |  | < 0.001 |
| Less than \$35,000 | 1,969 (29.21) | 468 (35.78) | 415 (46.16) | 111 (23.87) | 878 (22.83) | 97 (43.69) | |
| \$35,000 - \$49,999 | 753 (11.17) | 180 (13.76) | 111 (12.35) | 39 (8.39) | 394 (10.24) | 29 (13.06) | |
| \$50,000 - \$69,999 | 959 (14.23) | 210 (16.06) | 148 (16.46) | 58 (12.47) | 520 (13.52) | 23 (10.36) | |
| \$70,000 - \$99,999 | 1,058 (15.70) | 179 (13.69) | 82 (9.12) | 88 (18.92) | 683 (17.76) | 26 (11.71) | |
| \$100,000+ | 1,793 (26.60) | 228 (17.43) | 115 (12.79) | 142 (30.54) | 1,266 (32.92) | 42 (18.92) | |
| Don't know | 208 (3.09) | 43 (3.29) | 28 (3.11) | 27 (5.81) | 105 (2.73) | 5 (2.25) |  |
| **Any Children <18, n (%)** |  |  |  |  |  |  | < 0.001 |
| No | 4,564 (67.72) | 692 (52.91) | 534 (59.40) | 314 (67.53) | 2,879 (74.86) | 145 (65.32) |  |
| Yes | 2,176 (32.28) | 616 (47.09) | 365 (40.60) | 151 (32.47) | 967 (25.14) | 77 (34.68) |  |

For seroconversion analyses, we used a subset of 3,422 participants seronegative in May-September 2020 and testing during November 2020-January 2021 (subset of testers). Compared to the full cohort, the subset of testers had more White NH participants (57% versus 67%), was older (mean: 44 years versus 41 years) and had higher educational attainment (57% versus 67% with at least a college education) (Supplemental Table 1).

### Measures of Exposure: Structural and Work-Related Ability to Social Distance

For structural measures of exposure (Table 2), a greater proportion of Hispanic, Black NH and API NH participants lived in an urban area and lived in a multi-unit dwelling compared to White NH participants. A greater proportion of Hispanic and Black NH participants were unable to avoid public transportation compared to API NH and White NH participants. For work-related measures, the proportion of participants with less ability to social distance was generally highest among Black NH participants and lowest among White NH participants. A greater proportion of Black NH than White NH participants who were employed reported that they were unable to work from home and could lose their job if unable to go to work. All differences were statistically significant (p<0.001). The proportion with more exposure risk was highest among Black NH (51%) and Hispanic (46%) and lowest among API NH (36%) and White NH participants (33%) (p<0.001).

**Table 2.**
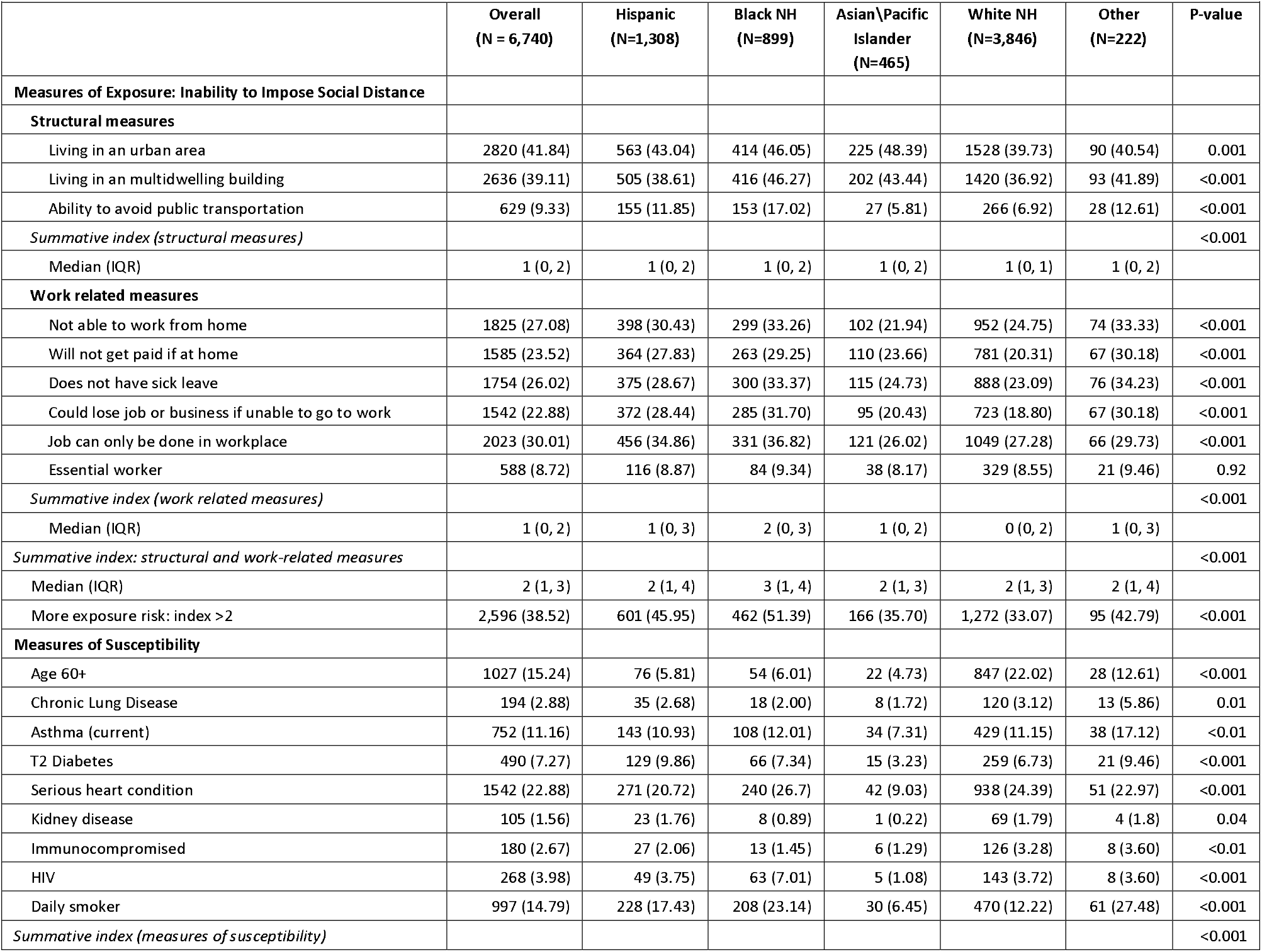
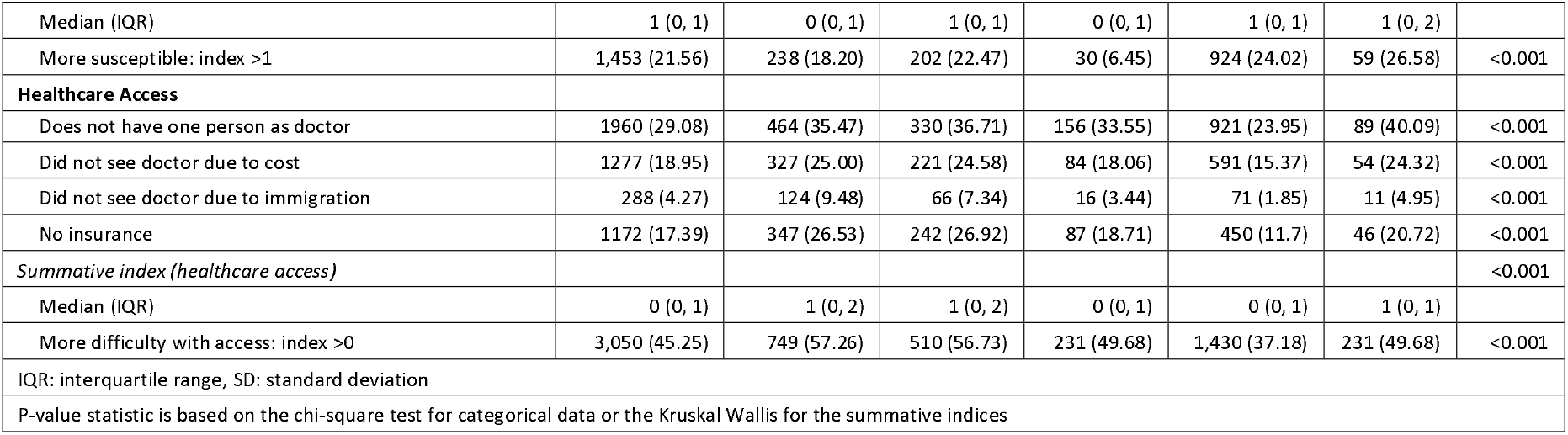
Measures of Exposure, Susceptibility and Access to Care - Stratified by Race/Ethnicity.

|  |  |  |  |  |  |  |  |
| --- | --- | --- | --- | --- | --- | --- | --- |
|  | Overall<br>(N = 6,740) | Hispanic<br>(N=1,308) | Black NH<br>(N=899) | Asian\Pacific<br>Islander<br>(N=465) | White NH<br>(N=3,846) | Other<br>(N=222) | P-value |
| **Measures of Exposure: Inability to Impose Social Distance** |  |  |  |  |  |  |  |
| **Structural measures** |  |  |  |  |  |  |  |
| Living in an urban area | 2820 (41.84) | 563 (43.04) | 414 (46.05) | 225 (48.39) | 1528 (39.73) | 90 (40.54) | 0.001 |
| Living in an multidwelling building | 2636 (39.11) | 505 (38.61) | 416 (46.27) | 202 (43.44) | 1420 (36.92) | 93 (41.89) | <0.001 |
| Ability to avoid public transportation | 629 (9.33) | 155 (11.85) | 153 (17.02) | 27 (5.81) | 266 (6.92) | 28 (12.61) | <0.001 |
| *Summative index (structural measures)* |  |  |  |  |  |  | <0.001 |
| Median (IQR) | 1 (0, 2) | 1 (0, 2) | 1 (0, 2) | 1 (0, 2) | 1 (0, 1) | 1 (0, 2) |  |
| **Work related measures** |  |  |  |  |  |  |  |
| Not able to work from home | 1825 (27.08) | 398 (30.43) | 299 (33.26) | 102 (21.94) | 952 (24.75) | 74 (33.33) | <0.001 |
| Will not get paid if at home | 1585 (23.52) | 364 (27.83) | 263 (29.25) | 110 (23.66) | 781 (20.31) | 67 (30.18) | <0.001 |
| Does not have sick leave | 1754 (26.02) | 375 (28.67) | 300 (33.37) | 115 (24.73) | 888 (23.09) | 76 (34.23) | <0.001 |
| Could lose job or business if unable to go to work | 1542 (22.88) | 372 (28.44) | 285 (31.70) | 95 (20.43) | 723 (18.80) | 67 (30.18) | <0.001 |
| Job can only be done in workplace | 2023 (30.01) | 456 (34.86) | 331 (36.82) | 121 (26.02) | 1049 (27.28) | 66 (29.73) | <0.001 |
| Essential worker | 588 (8.72) | 116 (8.87) | 84 (9.34) | 38 (8.17) | 329 (8.55) | 21 (9.46) | 0.92 |
| *Summative index (work related measures)* |  |  |  |  |  |  | <0.001 |
| Median (IQR) | 1 (0, 2) | 1 (0, 3) | 2 (0, 3) | 1 (0, 2) | 0 (0, 2) | 1 (0, 3) |  |
| *Summative index: structural and work-related measures* |  |  |  |  |  |  | <0.001 |
| Median (IQR) | 2 (1, 3) | 2 (1, 4) | 3 (1, 4) | 2 (1, 3) | 2 (1, 3) | 2 (1, 4) |  |
| More exposure risk: index >2 | 2,596 (38.52) | 601 (45.95) | 462 (51.39) | 166 (35.70) | 1,272 (33.07) | 95 (42.79) | <0.001 |
| **Measures of Susceptibility** |  |  |  |  |  |  |  |
| Age 60+ | 1027 (15.24) | 76 (5.81) | 54 (6.01) | 22 (4.73) | 847 (22.02) | 28 (12.61) | <0.001 |
| Chronic Lung Disease | 194 (2.88) | 35 (2.68) | 18 (2.00) | 8 (1.72) | 120 (3.12) | 13 (5.86) | 0.01 |
| Asthma (current) | 752 (11.16) | 143 (10.93) | 108 (12.01) | 34 (7.31) | 429 (11.15) | 38 (17.12) | <0.01 |
| T2 Diabetes | 490 (7.27) | 129 (9.86) | 66 (7.34) | 15 (3.23) | 259 (6.73) | 21 (9.46) | <0.001 |
| Serious heart condition | 1542 (22.88) | 271 (20.72) | 240 (26.7) | 42 (9.03) | 938 (24.39) | 51 (22.97) | <0.001 |
| Kidney disease | 105 (1.56) | 23 (1.76) | 8 (0.89) | 1 (0.22) | 69 (1.79) | 4 (1.8) | 0.04 |
| Immunocompromised | 180 (2.67) | 27 (2.06) | 13 (1.45) | 6 (1.29) | 126 (3.28) | 8 (3.60) | <0.01 |
| HIV | 268 (3.98) | 49 (3.75) | 63 (7.01) | 5 (1.08) | 143 (3.72) | 8 (3.60) | <0.001 |
| Daily smoker | 997 (14.79) | 228 (17.43) | 208 (23.14) | 30 (6.45) | 470 (12.22) | 61 (27.48) | <0.001 |
| *Summative index (measures of susceptibility)* |  |  |  |  |  |  | <0.001 |
| Median (IQR) | 1 (0, 1) | 0 (0, 1) | 1 (0, 1) | 0 (0, 1) | 1 (0, 1) | 1 (0, 2) |  |
| More susceptible: index >1 | 1,453 (21.56) | 238 (18.20) | 202 (22.47) | 30 (6.45) | 924 (24.02) | 59 (26.58) | <0.001 |
| **Healthcare Access** |  |  |  |  |  |  |  |
| Does not have one person as doctor | 1960 (29.08) | 464 (35.47) | 330 (36.71) | 156 (33.55) | 921 (23.95) | 89 (40.09) | <0.001 |
| Did not see doctor due to cost | 1277 (18.95) | 327 (25.00) | 221 (24.58) | 84 (18.06) | 591 (15.37) | 54 (24.32) | <0.001 |
| Did not see doctor due to immigration | 288 (4.27) | 124 (9.48) | 66 (7.34) | 16 (3.44) | 71 (1.85) | 11 (4.95) | <0.001 |
| No insurance | 1172 (17.39) | 347 (26.53) | 242 (26.92) | 87 (18.71) | 450 (11.7) | 46 (20.72) | <0.001 |
| *Summative index (healthcare access)* |  |  |  |  |  |  | <0.001 |
| Median (IQR) | 0 (0, 1) | 1 (0, 2) | 1 (0, 2) | 0 (0, 1) | 0 (0, 1) | 1 (0, 1) |  |
| More difficulty with access: index >0 | 3,050 (45.25) | 749 (57.26) | 510 (56.73) | 231 (49.68) | 1,430 (37.18) | 231 (49.68) | <0.001 |
| IQR: interquartile range, SD: standard deviation |  |  |  |  |  |  |  |
| P-value statistic is based on the chi-square test for categorical data or the Kruskal Wallis for the summative indices |  |  |  |  |  |  |  |

### Measures of Susceptibility

API NH participants, generally, had the lowest frequency of individual metrics of COVID-19 susceptibility. Hispanic, Black and White NH participants were more likely to report a serious heart condition and current asthma than API NH participants (p<0.01). Hispanic and Black NH participants were more likely to report daily smoking than API NH or White NH participants (p<0.001). The proportion more susceptible was higher among White NH (24%), Black NH (23%), and Hispanic (18%) participants than API NH participants (7%) (p<0.001).

### Measures of Healthcare Access

Hispanic, Black NH and API NH participants were more likely to report having no primary care doctor, not seeing a doctor due to cost, not seeing a doctor due to immigration, and not having insurance than White NH participants (p <0.001). The proportion with more difficulty with access to healthcare was higher among Hispanic (57%), Black NH (57%), and API NH participants (50%) than White NH participants (37%) (p<0.001).

Broadly, the trends in exposure, susceptibility and healthcare access in the subset of testers mirrored trends in the full cohort (Supplemental Table 2).

### Association of Exposure, Susceptibility, and Access to Care with COVID-19 Outcomes

Approximately 5% (N = 161/3422) seroconverted and 6% (N = 401/6070) were hospitalized (Table 3). In models adjusted for socio-demographics including age, participants with more (versus less) exposure risk had greater odds of seroconversion (aOR: 1.64, 95% CI: 1.18-2.29) and hospitalization (aOR: 1.73; 1.40-2.14) (Table 3). Neither susceptibility nor access to care was associated with seroconversion. However, participants with more (versus less) susceptibility and those with more (versus less) difficulty with healthcare access had greater odds of hospitalization (aOR_susceptibility_: 2.36; 1.90-2.96 and aOR_access_: 2.31; 1.69-2.68).

**Table 3.** Impact of Exposure, Susceptibility, and Access to Health Care on the Odds Seroconversion (N = 3,422) and Hospitalization (N = 6,070)

|  |  |  |  |  |  |  |
| --- | --- | --- | --- | --- | --- | --- |
|  | **Seroconversion** |  |  | **Hospitalization** |  |  |
|  | **N (%)** | **OR (95% CI)** | **aOR(95% CI)** | **N (%)** | **OR (95% CI)** | **aOR(95% CI)** |
| **Overall** | 161 (4.70) | 161 (4.70) | 161 (4.70) | 401 (5.95) |  |  |
| **Exposure (Structural + Work Related)<sup>1</sup>** |  |  |  |  |  |  |
| Less exposure risk | 86 (3.73) | 1.00 | 1.00 | 178 (4.30) | 1.00 | 1.00 |
| More exposure risk | 75 (6.73) | **1.86 (1.35, 2.56)** | **1.64 (1.18, 2.29)** | 223 (8.59) | **2.09 (1.71, 2.57)** | **1.73 (1.40, 2.14)** |
| **Susceptibility<sup>2</sup>** |  |  |  |  |  |  |
| Less susceptible | 130 (4.95) | 1.00 | 1.00 | 258 (4.88) | 1.00 | 1.00 |
| More susceptible | 31 (3.90) | 0.78 (0.52, 1.16) | 0.75 (0.50, 1.12) | 143 (9.84) | **2.13 (1.72, 2.63)** | **2.36 (1.90, 2.96)** |
| **Access to Healthcare<sup>1</sup>** |  |  |  |  |  |  |
| Less barriers to access | 93 (4.21) | 1.00 | 1.00 | 130 (3.52) | 1.00 | 1.00 |
| More barriers to access | 68 (5.61) | 1.35 (0.98, 1.86) | 1.21 (0.86, 1.69) | 271 (8.89) | **2.67 (2.15, 3.31)** | **2.13 (1.69, 2.68)** |
| aOR = Adjusted Odds Ratio, 95% CI = 95% Confidence Interval |  |  |  |  |  |  |
| 1. Model adjusted for race, gender, age, presence of children in the household, income, education, loss of income/employment. |  |  |  |  |  |  |
| 2. Model adjusted for race, gender, presence of children in the household, income, education, and loss of income/employment |  |  |  |  |  |  |

### EMM by Race/Ethnicity

Hispanics and Black NH were more likely to seroconvert or to be hospitalized for COVID-19 than API NH or White NH participants (seroconversion: 7%, 6%, versus 4%, 3%, respectively, p<0.01; hospitalization: 8%, 9% versus 5%, 3%, respectively, p<0.001) (Supplemental Table 3). For the seroconversion outcome, we saw no evidence of EMM by race/ethnicity (Supplemental Table 4). For the hospitalization outcome, we saw evidence of EMM by race/ethnicity for the susceptibility exposure (RERI: 1.07, p=0.03), meaning that non-White participants with a high score on the susceptibility index were at disproportionately higher odds of COVID hospitalization, compared with White participants. The odds of COVID hospitalization were 2.79 (2.06-3.78) among non-White participants and 2.04 (1.48-2.81) among White participants. In contrast, there was no evidence of EMM by race/ethnicity for the COVID-19 exposure or health care access indices with hospitalization (Table 4).

**Table 4.**
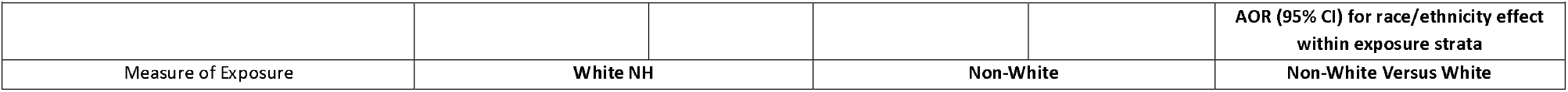
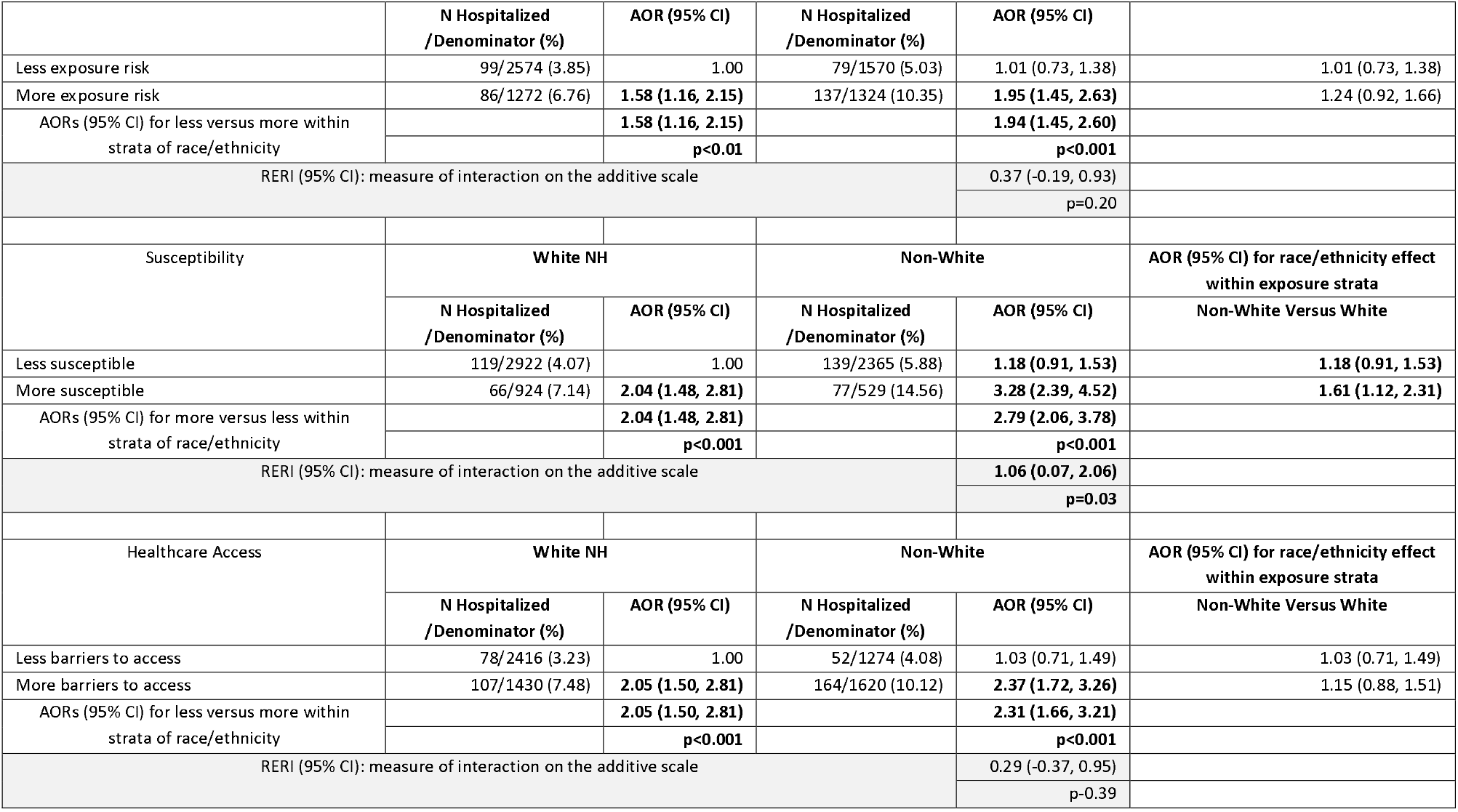
Modification of the association between race/ethnicity and hospitalization by (A) inability to impose social distance, (B) susceptibility and (C) healthcare access – N = 6,070.

## DISCUSSION

Our study confirms the existence of significant racial and ethnic differences in potential COVID-19 exposure risk, susceptibility to COVID-19 complications, and access to health care within a large U.S. national cohort. The proportion with more exposure risk and more difficulty with healthcare access was significantly higher among Black NH, Hispanic and API NH than White NH participants. More exposure, as measured by reduced ability to social distance, increased the odds of seroconversion by 64% and hospitalization by 73%. More underlying susceptibility and more difficulty with access to care increased the odds of hospitalization by 113-136%.

Many have hypothesized that SEP influences the differential impact of the COVID-19 pandemic either directly or indirectly via occupation, living and working conditions, health-related behaviors, presence of comorbidities and immune functioning.(8,11,12) However, the influence of SEP on COVID-19 transmission, severity and outcomes is understudied, and existing research has largely characterized SEP using geography and race/ethnicity as a proxies.(21–27) For example, U.S. counties with a higher proportion of Black or Hispanic population or of adults with less than a high school diploma had disproportionately higher COVID-19 cases.(26) Using data from the American Community Survey to characterize socioeconomic vulnerability at the neighborhood level, ecologic analyses have demonstrated that increasing levels of socioeconomic vulnerability were associated with gaps in COVID-19 testing coverage in Massachusetts and COVID-19 mortality in Chicago.(21,27) Although useful, such approaches may mask the extent of inequity and the influence of SEP at the individual-level. We are aware of one study that included individual-level social indicators to assess COVD-19 outcomes.(28) Hispanic ethnicity, inability to shelter in place and maintain income, frontline service work, unemployment, and household income <$50,000 increased the risk of COVID-19 infection among residents and workers located in small community within San Francisco.(28) We provide empirical evidence to support the conceptual model of Blumenshine, et al, in the context of the COVID-19 pandemic.(12) Differences in SEP contribute to disparities in exposure, susceptibility to illness given infection, and timely access to care and supportive treatment. Further, reduced ability to social distance was positively associated with seroconversion and hospitalization, and increased susceptibility to COVID-19 complications and poor access to healthcare were positively associated with hospitalization. We did not observe an association between seroconversion with susceptibility or access to care. The null finding is unsurprising given susceptibility to complications and access to care would be expected to influence illness following infection. Primary and secondary prevention efforts should address potential social disparities in exposure, COVID-19 vaccination, and access to care/treatment.

Our finding that Hispanic and Black NH participants had more exposure and more difficulty with healthcare access than White NH participants is consistent with other research showing a disproportionate burden of COVID-19 infections, complications and mortality on racial and ethnic minorities.(3,5,8,23,29–35) The positive additive interaction observed between racial and ethnic minority group status and susceptibility to severe COVID-19 disease with hospitalization is especially concerning. We did not observe evidence of EMM by race/ethnicity in terms of the COVID exposure index or the healthcare access index. The implication of these findings is that non-White populations should be prioritized in efforts to reduce susceptibility, whereas interventions to reduce COVID exposure (i.e., social distancing ability) and healthcare access should target everyone. Recommendations for and discussions about social distancing fail to account for the reality of differential ability to adopt and benefit from these approaches, creating inequities in health outcomes. Long-standing health disparities contribute to susceptibility among Hispanic and Black NH individuals and susceptibility is also influenced by lower healthcare access. Mitigation strategies and messaging should intensify focus on Hispanic and Black NH individuals with conditions that increase risk of COVID-19 morbidity and mortality and incorporate targeted, culturally appropriate communication.

As with all observational studies, confounding of the exposure-outcome relationship is a concern, and we did not address the possibility of joint effects of the SEP indices. All exposure data was collected at cohort enrollment. The benefit of using enrollment data for exposure measurement is that all participants had to complete the screening tool to enroll into prospective follow-up. Thus, missing data was minimized. Measurement error may be a concern given data on hospitalizations was self-reported.

There have been increasing calls for research to better capture and report on socioeconomic determinants of COVID-19 outcomes alongside race/ethnicity to identify populations that may experience a disproportionate burden of risk and/or ability to benefit from pandemic mitigation strategies.(10,13) We observed significant racial/ethnic disparities in ability to social distance as a measure of COVID exposure, susceptibility to COVID-19 complications and access to healthcare in our national cohort. To the best of our knowledge, we present some of the first data, measured at the individual level, to demonstrate racial/ethnic differences in these factors and their association with seroconversion and hospitalization. Future pandemic mitigation strategies should account for the inequities in burden that can be introduced via SEP in relation to pathogen exposure, susceptibility, and health care access.

## Data Availability

All data produced in the present study are available upon reasonable request to the authors

https://cunyisph.org/chasing-covid/

## Abbreviations (in order of appearance)

introduction
(COVID-19): Coronavirus Disease 2019
(CDC): Centers for Disease Control and Prevention
(SEP): socioeconomic position
(CLI): COVID-like illness
(EMM): effect measure modification
methods
(US): United States
(NH): Non-Hispanic
(API): Asian/Pacific Islander
(CSTE): Council of State and Territorial Epidemiologists
(RERI): relative excess risk due to interaction

## CONTRIBUTORS

MMR and DN conceptualized the study. MMR and MS performed statistical analyses. MMR and MS wrote the first draft of the paper. MMR, EB, DN, and MS contributed to interpreting the data. MMR, MS, EB, DN, MM, MR DAW, AMP, RZ, ARM, SGK, and CG contributed to the writing and revising of the manuscript. MR, MMR, SGK and DN contributed to data collection, cleaning and management. DN, SGK, MMR and CG contributed to obtaining funding for the research.

## ACKNOWLEDGEMENTS

The authors wish to thank the participants of the CHASING COVID Cohort Study. We are grateful to you for your contributions to the advancement of science around the SARS-CoV-2 pandemic.

---

**Supplemental Table 1.** Demographic and Socioeconomic Characteristics of the Chasing COVID Cohort (C3) Participants who were Seronegative and Re-Tested - United States, Enrolled from March 28 - April 20, 2020 (N = 3,422)

|  | Total | Hispanic | Black NH | Asian\Pacific<br>Islander NH | White NH | Other NH | P-value |
| --- | --- | --- | --- | --- | --- | --- | --- |
| **Total, n (%)** | 3,422 (100.00) | 500 (14.61) | 279 (8.15) | 219 (6.4) | 2,312 (67.56) | 112 (3.28) |  |
| **Age** |  |  |  |  |  |  | <<br>0.001 |
| Mean (SD) | 44 (15.16) | 39.51 (13.13) | 40.5 (12.90) | 33 (13.18) | 46.62 (15.43) | 44.33 (13.57) |  |
| Median (IQR) | 42 (32, 56) | 37 (30, 48) | 37 (30, 51) | 33 (26, 41) | 45 (34, 60) | 43 (35, 53) |  |
| **Age Category, n (%)** |  |  |  |  |  |  | <<br>0.001 |
| 18-29 | 617 (18.03) | 120 (24.00) | 57 (20.43) | 84 (38.36) | 342 (14.79) | 14 (12.50) |  |
| 30-39 | 934 (27.29) | 166 (33.20) | 100 (35.84) | 76 (34.70) | 566 (24.48) | 26 (23.21) |  |
| 40-49 | 654 (19.11) | 104 (20.80) | 50 (17.92) | 30 (13.70) | 430 (18.60) | 40 (35.71) |  |
| 50-59 | 514 (15.02) | 62 (12.40) | 40 (14.34) | 11 (5.02) | 388 (16.78) | 13 (11.61) |  |
| 60+ | 703 (20.54) | 48 (9.60) | 32 (11.47) | 18 (8.22) | 586 (25.35) | 19 (16.96) |  |
| **Gender, n (%)** |  |  |  |  |  |  | <<br>0.001 |
| Male | 1,516 (44.30) | 219 (43.80) | 124 (44.44) | 88 (40.18) | 1,033 (44.68) | 52 (46.43) |  |
| Female | 1,810 (52.89) | 267 (53.40) | 147 (52.69) | 128 (58.45) | 1,221 (52.81) | 47 (41.96) |  |
| Non- Binary | 96 (2.81) | 14 (2.80) | 8 (2.87) | 3 (1.37) | 58 (2.51) | 13 (11.61) |  |
| **Education, n (%)** |  |  |  |  |  |  | <<br>0.001 |
| Less than 12th grade | 37 (1.08) | 10 (2.00) | 2 (0.72) | 2 (0.91) | 22 (0.95) | 1 (0.89) |  |
| 12th grade/GED | 278 (8.12) | 71 (14.20) | 33 (11.83) | 6 (2.74) | 155 (6.70) | 13 (11.61) |  |
| Some college (1-3 years) | 815 (23.82) | 151 (30.20) | 112 (40.14) | 38 (17.35) | 488 (21.11) | 26 (23.21) |  |
| College (4 or more years) | 2,292 (66.98) | 268 (53.60) | 132 (47.31) | 173 (79.00) | 1,647 (71.24) | 72 (64.29) |  |
| **Employment Status, n (%)** |  |  |  |  |  |  | <<br>0.001 |
| Employed | 2,102 (61.43) | 284 (56.80) | 181 (64.87) | 131 (59.82) | 1,436 (62.11) | 70 (62.50) |  |
| Out of Work | 402 (11.75) | 88 (17.60) | 42 (15.05) | 26 (11.87) | 229 (9.90) | 17 (15.18) |  |
| Other | 918 (26.83) | 128 (25.60) | 56 (20.07) | 62 (28.31) | 647 (27.98) | 25 (22.32) |  |
| **Income, n (%)** |  |  |  |  |  |  | <<br>0.001 |
| Less than \$35,000 | 870 (25.42) | 174 (34.80) | 113 (40.50) | 47 (21.46) | 494 (21.37) | 42 (37.50) | |
| \$35,000 - \$49,999 | 383 (11.19) | 77 (15.40) | 43 (15.41) | 16 (7.31) | 231 (9.99) | 16 (14.29) | |
| \$50,000 - \$69,999 | 505 (14.76) | 75 (15.00) | 56 (20.07) | 30 (13.70) | 333 (14.40) | 11 (9.82) | |
| \$70,000 - \$99,999 | 592 (17.30) | 73 (14.60) | 30 (10.75) | 43 (19.63) | 436 (18.86) | 10 (8.93) | |
| \$100,000+ | 993 (29.02) | 90 (18.00) | 33 (11.83) | 72 (32.88) | 768 (33.22) | 30 (26.79) | |
| Don't know | 79 (2.31) | 11 (2.20) | 4 (1.43) | 11 (5.02) | 50 (2.16) | 3 (2.68) |  |
| **Any Children <18, n (%)** |  |  |  |  |  |  | <<br>0.001 |
| No | 2,596 (75.86) | 326 (65.20) | 183 (65.59) | 154 (70.32) | 1,853 (80.15) | 80 (71.43) |  |
| Yes | 826 (24.14) | 174 (34.80) | 96 (34.41) | 65 (29.68) | 459 (19.85) | 32 (28.57) |  |

IQR: Interquartile Range, NH: non-Hispanic, n: number, SD: standard deviation, US: United States, and %: percentage
\*P-value based on the kruskall wallis test for means and Chi Square for frequencies

**Supplemental Table 2.** Measures of Exposure, Susceptibility and Access to Care – Among Participants who were Seronegative and Re-Tested (N = 3,422)

|  |  |  |  |  |  |  |  |
| --- | --- | --- | --- | --- | --- | --- | --- |
|  | Overall<br>(N = 6,740) | Hispanic<br>(N=1,308) | Black NH<br>(N=899) | Asian\Pacific<br>Islander<br>(N=465) | White NH<br>(N=3,846) | Other<br>(N=222) | P-value |
| **Measures of Exposure: Inability to Impose Social Distance** |  |  |  |  |  |  |  |
| **Structural measures** |  |  |  |  |  |  |  |
| Living in an urban area | 1,430 (41.79) | 219 (43.80) | 140 (50.18) | 116 (52.97) | 906 (39.19) | 49 (43.75) | <0.001 |
| Living in an multidwelling building | 1,348 (39.39) | 206 (41.20) | 144 (51.61) | 93 (42.47) | 859 (37.15) | 46 (41.07) | <0.001 |
| Ability to avoid public transportation | 218 (6.37) | 38 (7.60) | 37 (13.26) | 7 (3.20) | 128 (5.54) | 8 (7.14) | <0.001 |
| *Summative index (structural measures)* |  |  |  |  |  |  |  |
| Median (IQR) | 1 (0,2) | 1 (0,2) | 1 (0,2) | 1 (0,2) | 1 (0,1) | 1 (0,2) | <0.001 |
| **Work related measures** |  |  |  |  |  |  |  |
| Not able to work from home | 849 (24.81) | 134 (26.80) | 87 (31.18) | 42 (19.18) | 554 (23.96) | 32 (28.57) | 0.01 |
| Will not get paid if at home | 632 (18.47) | 104 (20.80) | 70 (25.09) | 38 (17.35) | 398 (17.21) | 22 (19.64) | 0.01 |
| Does not have sick leave | 746 (21.80) | 116 (23.20) | 82 (29.39) | 46 (21.00) | 474 (20.50) | 28 (25.00) | 0.01 |
| Could lose job or business if unable to go to work | 597 (17.45) | 113 (22.60) | 69 (24.73) | 38 (17.35) | 353 (15.27) | 24 (21.43) | <0.001 |
| Job can only be done in workplace | 911 (26.62) | 157 (31.40) | 98 (35.13) | 50 (22.83) | 577 (24.96) | 29 (25.89) | <0.01 |
| Essential worker | 282 (8.24) | 34 (6.80) | 32 (11.47) | 15 (6.85) | 189 (8.17) | 12 (10.71) | 0.15 |
| *Summative index (work related measures)* |  |  |  |  |  |  | <0.001 |
| Median (IQR) | 1 (0,2) | 1 (0,3) | 2 (0,3) | 1 (0,2) | 0 (0,2) | 1 (0,3) |  |
| *Summative index: structural and work-related measures* |  |  |  |  |  |  |  |
| Median (IQR) | 2 (1,3) | 2 (1,4) | 3 (1,4) | 2 (1,3) | 2 (1,3) | 2 (1,4) | <0.001 |
| More exposure risk: index >2 | 1,115 (32.58) | 189 (37.80) | 130 (46.59) | 69 (31.51) | 687 (29.71) | 40 (35.71) | <0.001 |
| **Measures of Susceptibility** |  |  |  |  |  |  |  |
| Age 60+ | 703 (20.54) | 48 (9.60) | 32 (11.47) | 18 (8.22) | 586 (25.35) | 19 (16.96) | <0.001 |
| Chronic Lung Disease | 103 (3.01) | 17 (3.40) | 9 (3.23) | 5 (2.28) | 64 (2.77) | 8 (7.14) | 0.10 |
| Asthma (current) | 389 (11.37) | 62 (12.40) | 35 (12.54) | 10 (4.57) | 261 (11.29) | 21 (18.75) | <0.01 |
| T2 Diabetes | 231 (6.75) | 44 (8.80) | 29 (10.39) | 5 (2.28) | 140 (6.06) | 13 (11.61) | <0.01 |
| Serious heart condition | 865 (25.28) | 115 (23.00) | 89 (31.90) | 20 (9.13) | 613 (26.51) | 28 (25.00) | <0.001 |
| Kidney disease | 50 (1.46) | 7 (1.40) | 2 (0.72) | 1 (0.46) | 39 (1.69) | 1 (0.89) | 0.45 |
| Immunocompromised | 108 (3.16) | 13 (2.60) | 7 (2.51) | 3 (1.37) | 80 (3.46) | 5 (4.46) | 0.34 |
| HIV | 161 (4.70) | 25 (5.00) | 30 (10.75) | 3 (1.37) | 97 (4.20) | 6 (5.36) | <0.001 |
| Daily smoker | 343 (10.02) | 58 (11.60) | 49 (17.56) | 8 (3.65) | 202 (8.74) | 26 (23.21) | <0.001 |
| *Summative index (measures of susceptibility)* |  |  |  |  |  |  |  |
| Median (IQR) | 1 (0, 1) | 0 (0,1) | 1 (0,1) | 0 (0,1) | 1 (0,1) | 1 (0,2) | <0.001 |
| More susceptible: index >1 | 795 (23.23) | 98 (19.60) | 78 (27.96) | 15 (6.85) | 570 (24.65) | 34 (30.36) | <0.001 |
| **Healthcare Access** |  |  |  |  |  |  |  |
| Does not have one person as doctor | 808 (23.61) | 145 (29.00) | 74 (26.52) | 69 (31.51) | 483 (20.89) | 37 (33.04) | <0.001 |
| Did not see doctor due to cost | 510 (14.90) | 94 (18.80) | 58 (20.79) | 41 (18.72) | 295 (12.76) | 22 (19.64) | <0.001 |
| Did not see doctor due to immigration | 27 (0.79) | 13 (2.60) | 4 (1.43) | 5 (2.28) | 2 (0.09) | 3 (2.68) | <0.001 |
| No insurance | 384 (11.22) | 88 (17.60) | 56 (20.07) | 27 (12.33) | 195 (8.43) | 18 (16.07) | <0.001 |
| *Summative index (healthcare access)* |  |  |  |  |  |  |  |
| Median (IQR) | 0 (0,1) | 1 (0,2) | 1 (0,2) | 0 (0,1) | 0 (0,1) | 1 (0,1) | <0.001 |
| More difficulty with access: index >0 | 1,213 (35.45) | 217 (43.40) | 121 (43.37) | 97 (44.29) | 726 (31.40) | 52 (46.43) | <0.001 |
| IQR: interquartile range, SD: standard deviation |  |  |  |  |  |  |  |
| P-value statistic is based on the chi-square test for categorical data or the Kruskal Wallis for the summative indices |  |  |  |  |  |  |  |

**Supplemental Table 3.**
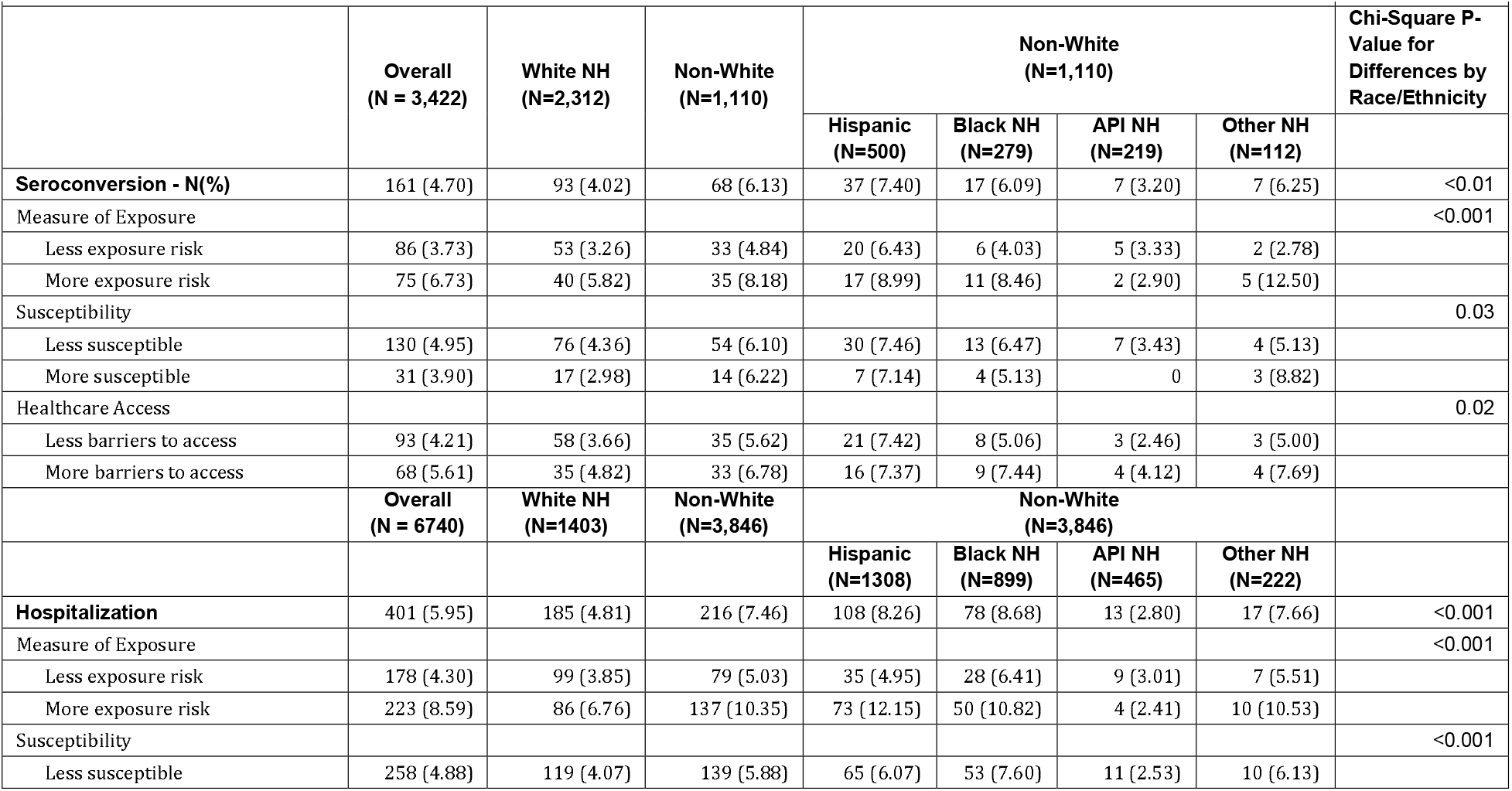
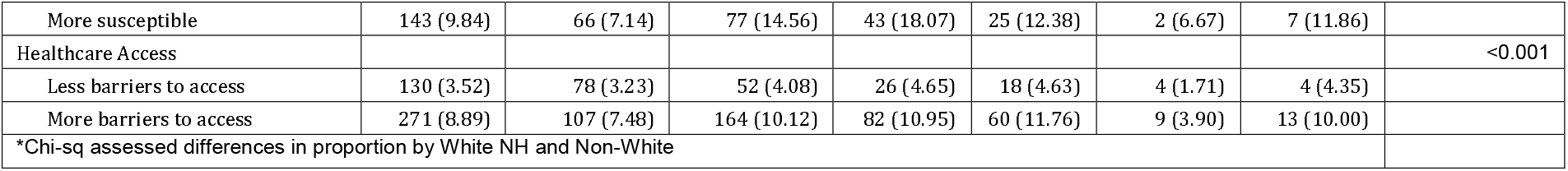
Proportion Hospitalized (Yes) or Seroconverted (Yes) By Exposure Level Within Race/Ethnicity Strata.

**Supplemental Table 4.**
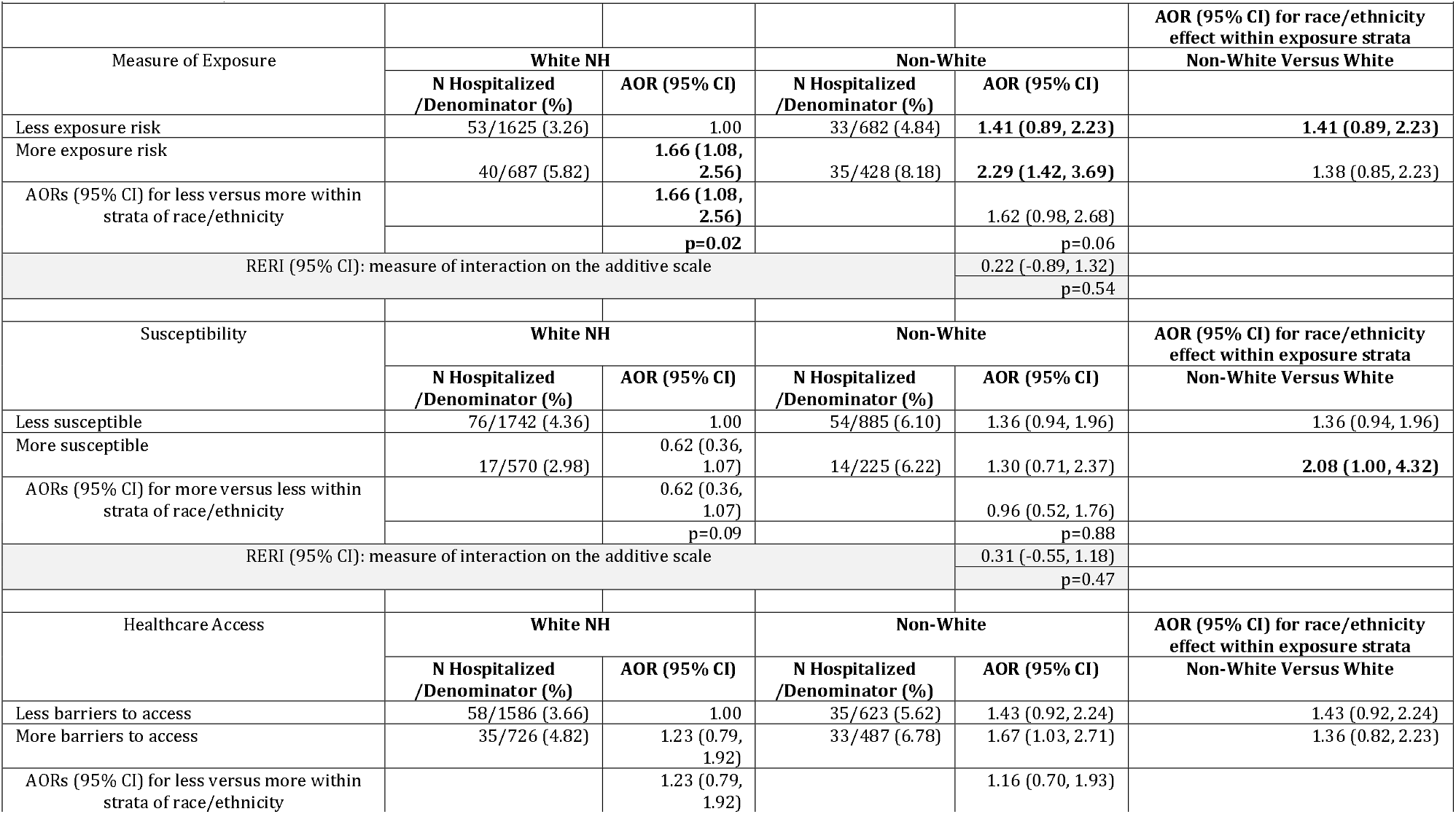
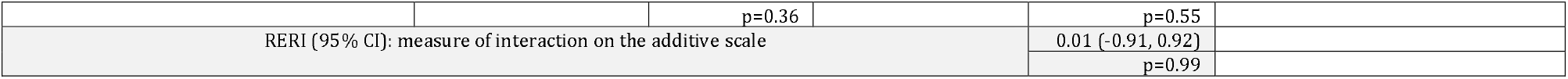
Modification of the association between race/ethnicity and seroconversion by (A) inability to impose social distance, (B) susceptibility and (C) healthcare access – N = 3,422.

